# Study protocol for The Heart Watch Study - Prognostic performance of smartwatch 12-lead ECG with advanced ECG analysis in consumer self-screening for cardiovascular disease

**DOI:** 10.1101/2025.03.05.25323393

**Authors:** Zaidon S Al-Falahi, Todd T Schlegel, Vivian Lee, Clara K Chow, Rebecca Kozor, Thomas Lindow, Martin Ugander

**Author notes:** denotes corresponding author.

## Abstract

**Background:** It is now possible to acquire a fully diagnostic standard 12-lead electrocardiography (ECG) using a smartwatch, a smartphone app, and no additional hardware. This study aims to evaluate the predictive performance of advanced ECG (A-ECG) analysis applied to a smartwatch 12-lead ECG (SWECG) acquired using an Apple Watch in identifying cardiovascular risks among asymptomatic adults performing consumer self-screening in the general population.

**Methods and Design:** The Heart Watch Study is a prospective, Australia-wide observational cohort study. The study will recruit 30,000 participants aged 20-79 years without prior known cardiovascular disease. Participants will enrol and consent electronically, and download a dedicated study iPhone app that enables a directed 12-lead SWECG acquisition using an Apple Watch. The recordings will undergo both conventional and A-ECG analysis. Follow-up will be performed through linkage with administrative health datasets in Australia, including hospital visits and mortality. The primary combined endpoint is all-cause mortality, hospitalization for cardiovascular causes, and incident cardiovascular disease (arrythmia, ischemic heart disease, heart failure) as predicted by existing A-ECG analyses.

**Conclusion:** This study evaluates the feasibility of performing a fully diagnostic standard 12-lead SWECG using only a smartwatch and a smartphone application, and addresses the prognostic value of A-ECG machine-learning based analysis in consumer self-screening in a large, diverse and apparently healthy population.

## Background

### Advanced ECG analysis of smartwatch 12-lead ECGs

Cardiovascular disease (CVD) benefits from early detection and intervention, yet presymptomatic features are difficult to identify, can often go unnoticed, and first presentation can be with an adverse event. The standard 12-lead ECG is a cornerstone of cardiac diagnostics worldwide for millions of patients every day ^1^. However, population screening for CVD using conventional ECG acquisition and interpretation is currently not recommended due to its cost and suboptimal sensitivity and specificity for pre-symptomatic disease ^2^. Our group has developed a validated method for recording accurate standard 12-lead ECG recordings using a smartwatch and a dedicated smartphone app with no additional hardware ^3, 4^. Furthermore, by incorporating advanced ECG analysis (A-ECG) of vectorcardiographic and waveform complexity measures derived from the standard 12-lead ECG, the accuracy of ECG for detecting CVD can be substantially improved ^5–8^.

### Rationale for Current Study

Coupling the innovations of A-ECG analysis with 12-lead ECG acquisition using increasingly ubiquitous consumer single-lead ECG technology such as smartwatches could overcome both the access and expertise barriers that currently limit ECG as a viable population-level screening tool. Attempts have been previously made to use smartphones or smartwatches to obtain a multi-lead ECG through either sequential recording or via interpolation using deep neural networks, but the resultant ECGs are fundamentally not identical to the standard 12-lead ECG ^8–10^. We have developed a method for calculation of all leads present in the standard 12-lead ECG by accurately time-aligning sequentially recorded channels. This allows for the correct calculation of Wilson’s central terminal that is critical for accurate derivation of the precordial leads. Taken together, this new 12-lead smartwatch ECG (SWECG) recording approach preserves the accuracy of the standard 12-lead ECG.

However, before clinical implementation, the association between pathological findings by 12-lead SWECG with A-ECG analysis and CVD outcomes remains to be determined.

In addition, a possible dose-response relationship between activity levels and health outcomes as suggested by the World Health Organization (WHO) is also in need of further research ^11^. The Heart Watch Study will also contribute to this research area by providing activity data from Apple Health paired with A-ECG analysis and long-term outcomes.

Finally, The A-ECG Heart Age is a prognostic metric of accelerated cardiovascular ageing that can be intuitively communicated to the public, potentially providing strong incentive for lifestyle interventions ^12, 13^. A-ECG Heart Age will be calculated from the 12-lead SWECG for all participants and validated against health outcomes. The results will be used to design further interventional studies to potentially slow down or reverse accelerated cardiovascular ageing.

## Objectives

### Primary Objective

To prospectively assess the prognostic performance of A-ECG risk scores from 12-lead SWECG for predicting cardiovascular events at 2 years in a broad and asymptomatic consumer self-screening population.

### Secondary Objectives

1. **Prognostic value of A-ECG Heart Age**: To examine the association of A-ECG Heart Age derived from the 12-lead SWECG, comparing it to age and health markers for cardiovascular risk prediction.
2. **Physical activity and cardiovascular health**: To examine the association of objective measures of physical activity from a mobile health device and CVRFs and future CVD events.
3. **Changes in A-ECG and physical activity**: To determine whether changes in physical activity over time correlate with changes in A-ECG Heart Age, providing insights into dynamic health status influenced by lifestyle factors.
4. **Real-world efficacy of A-ECG:** To evaluate the real-world diagnostic efficacy of A-ECG analysis of SWECG in a consumer self-screening setting.
5. **Screening and referral efficacy**: To assess the proportion of the study population that exhibits ECG and A-ECG findings necessitating a recommendation for further medical evaluation, particularly for relatively rare cardiac conditions, e.g., genetic ion channelopathies and cardiomyopathies, in an asymptomatic screening context.
6. **Long-term outcomes of A-ECG and activity scores**: To evaluate the long-term health outcomes associated with A-ECG scores and physical activity metrics beyond the initial two-year follow-up period.
7. **Compliance with WHO physical activity guidelines**: To quantify the proportion of the study population adhering to the World Health Organization’s physical activity guidelines and to examine the association of compliance with cardiovascular health outcomes.

## Methods

### Study design

A prospective, large-scale, observational cohort study with outcome ascertainment at 2 years via routinely collected health administrative datasets in Australia. The study is registered at anzctr.org.au (Australian New Zealand Clinical Trials Registry) under the identifier ACTRN12625000768493. Study recruitment will be via the website http://heartwatchstudy.com.au.

The study aims to recruit an age- and sex-stratified cohort of 30,000 participants (2500 per age and sex category) throughout Australia. No physical visits are required, enrolment and data collection are conducted entirely online.

### Eligibility

Eligibly participants will be those that meet all inclusion and exclusion criteria and provide electronic informed consent.

### Inclusion Criteria

Asymptomatic individuals 20 to 79 years of age without known CVD. Access to an ECG-enabled Apple Watch and a paired compatible iPhone.

### Exclusion Criteria

Exclusion criteria include Known heart disease including arrhythmia, cardiomyopathy, ischemic heart disease, congenital heart disease, previous cardiac interventions, e.g. percutaneous coronary intervention, coronary artery by-pass grafting, implanted cardiac devices, and bundle branch blocks, as well as inability or unwillingness to provide consent.

### Recruitment

Participants will be recruited from the general population through media campaigns (social and conventional media). Participants will complete an online screening questionnaire and provide baseline information on age, sex, contact information, and medical history.

Participants meeting eligibility criteria will then be issued a unique identification code to activate the free study smartphone app downloadable from the Apple App Store. The app will enable the participants to self-record the smartwatch 12-lead ECG, guiding them through the process, then transmitting it, along with Apple Health data, to secure research servers.

### Data Collection

In addition to the SWECG, physical activity and Apple Health data from the preceding 12 months will be extracted. All SWECGs will undergo manual ECG interpretation by cardiac technicians supervised by a board-certified cardiologist. Pre-determined criteria for triggering referral for further medical evaluation based on traditional 12-lead ECG findings will rely on international recommendations for ECG interpretation in athletes (Table 1) ^14^. Subsequently, all participants, including those meeting criteria for further medical evaluation, will undergo A-ECG analysis.

**Table 1.** Smartwatch 12-lead ECG findings to trigger referral to the primary care doctor.

| ECG abnormality | Definition |
| --- | --- |
| T wave inversion | $\geq 1$ mm in depth in two or more contiguous leads; excludes leads aVR, III and V1 |
| Anterior | V2-V4(excluding black athletes with J-point elevation and convex ST segment elevation followed by T-wave inversion in V2-V4; athletes < age 16 with T-wave inversion in V1-V3; and biphasic T waves in V3 only) |
| Lateral | I and aVL, V5 and/or V6 (only one lead of TWI required in V5 or V6) |
| Inferolateral | II and aVF, V5-V6, I and aVL |
| Inferolateral<br>Inferior | II and aVF, V5-V6, I and aVL<br>II and aVF |
| Inferior ST segment depression | II and aVF<br>$\geq 0.5$ mm in depth in two or more contiguous leads |
| Pathological Q waves | Q/R ratio $\geq 0.25$ or $\geq 40$ ms in duration in two or more leads (excluding III and aVR) |
| Complete left bundle branch block | QRS $\geq 120$ ms, predominantly negative QRS complex in lead V1 (QS or rS) and upright notched or slurred R wave in leads I and V6 |
| Non-specific intraventricular conduction delay | Any QRS duration $\geq 140$ ms |
| Epsilon wave | Distinct low amplitude signal (small positive deflection or notch) between the end of the QRS complex and onset of the T wave in leads V1-V3 |
| Ventricular pre-excitation | PR interval <120 ms with a delta wave (slurred upstroke in the QRS complex) and wide QRS ($\geq 120$ ms) |
| Prolonged QT interval* | QTc $\geq 470$ ms (male) |
| Prolonged QT interval* | QTc $\geq 480$ ms (female) |
| Brugada type 1 pattern | QTc ≥500 ms (marked QT prolongation) |
| | Coved pattern: initial ST elevation ≥2 mm (high take-off) with downsloping ST segment elevation followed by a negative symmetric T wave in ≥1 leads in V1-V3 |
| Profound sinus bradycardia | $<$30 beats per minute or sinus pauses ≥3 s |
| Profound 1° atrioventricular block | ≥400 ms |
| Mobitz type II 2° atrioventricular block | Intermittently non-conducted P waves with a fixed PR interval |
| 3° atrioventricular block | Complete heart block |
| Atrial tachyarrhythmias | Supraventricular tachycardia ($>$100bpm), atrial fibrillation, atrial flutter |
| Premature ventricular ectopic complexes | ≥2 premature ventricular ectopic complexes per 10 s tracing |
| Ventricular arrhythmias | Couplets, triplets and non-sustained ventricular tachycardia (frequent) |

### Study duration and follow-up

Participants are required to only record the initial enrolment SWECG. Two years after recruitment, participant data will be linked with data from national administrative health linkage datasets, which includes incident major cardiovascular diagnoses, all-cause hospitalisation, and death, including cause of death.

A subset of participants (n=1000) will be contacted electronically (by SMS and/or email) after 12 months, and asked to perform an additional 12-lead SWECG recording, and to share physical activity data of the preceding 12 months. This subset will be selected based on age, sex, A-ECG Heart Age, availability of physical activity levels in the initial extraction, and the quality of the 12-lead ECG recording. A-ECG Heart Age is a validated estimate of accelerated cardiovascular aging expressed in years of healthy human ageing, while the Heart Age gap is the difference between Heart Age and chronological age ^12, 13^. This sub-analysis will provide information on changes in ECG Heart age and physical activity levels.

### Collection of Apple Health Information

For purpose of the study, the study app will request to read and share permissions that the participant can individually authorize or reject as follows:

- **Activity data** including steps, walking and running distance, active energy, resting energy, standing time, exercise time, flights climbed, workouts, derived cardio fitness in VO2 max, cycling distance, swimming distance and strokes, wheelchair pushes and distance for wheelchair-bound individuals if applicable.
- **Body measurements including** height, weight, body mass index (BMI), body fat percentage, lean body mass, waist circumference, body temperature, and electrodermal activity.
- **Cardiac data** including mean heart rate, heart rate at rest, and walking or exercising, heart rate variability, ECGs, blood pressure measurements, and notifications about low, high and irregular heart rate.
- **Mobility data** including double support time, step length, walking asymmetry, walking speed, walking steadiness, six-minute walk distance, speed on stairs, falls and notifications.
- **Sleep data** including sleep patterns, schedule, and hours.

### Hardware and Software Versions

Access to An ECG-enabled Apple Watch Series 4 or later (not Apple Watch SE) and a paired iPhone are required for enrolment. A person can be enrolled using the watch and phone of someone else who is willing to help them participate in the study. Health data from the devices that such people use to enrol will not be included for analysis. The study app tracks the iPhone and Apple Watch versions as well as the iOS and WatchOS versions to enable analysis of the potential impact on ECG data of the given software version, if/when relevant.

### Study visits and procedures schedule

The study does not include any in-person visits for study participants. Recruitment and consent will be performed entirely online, and participants will only be contacted in the following circumstances:

1. Detection of a major ECG abnormality according to pre-set criteria on their 12-lead ECG (Table 1). The participant will then receive written information with the recommendation to seek evaluation from a primary care doctor as detailed further below.
2. Possible request for a repeat ECG recording if initial ECG was unsuitable for analysis due to poor quality, significant artifacts, or incomplete/incorrect recording.
3. Contact to request participation in a follow-up study at 12 months for another self-recorded ECG recording and extraction of activity data as described earlier.

All contact will be written via SMS and/or email to the contact information supplied by the participant at enrolment.

### Potential study risks

The activity of recording an ECG is minimal risk, and No adverse events are expected from self-recording of ECGs. Participating in a study aiming to identify people at risk of CVD may lead to a misconception from the participant that they are being monitored for disease, which may affect their decision on whether to seek medical assistance in the occurrence of cardiac symptoms. This risk will be reduced by thorough participant information to the study participants.

Data research and collection of personal information is inherently associated with risk of spread of personal information should a security breach occur. Data collected from participants will be stored in University of Sydney’s dedicated and secure Research Electronic Data Capture (REDCap) database. Anonymized A-ECG data will be securely stored using University of Sydney’s dedicated Amazon Web Services cloud infrastructure.

### ECG abnormalities to potentially trigger further evaluation

This is an observational cohort study to assess the prognostic performance of novel technology (A-ECG analysis of 12-lead SWECG) to detect subclinical CVD in a population without prior known heart disease. There is no available evidence to support clinical decision making in such a circumstance.

This study does not interfere with the current standards of care. The value of incidental detection of ECG abnormalities requiring further clinical work up on screening of an apparently healthy population is a matter of ongoing research ^15–19^.

In an Australian validation of an earlier version of the international consensus (Seattle criteria), out of 1197 Australian athletes screened, 4.5% were found to have abnormalities and only 0.3% had true underlying life-threatening abnormalities, yielding a false positive rate of 4.2%. The International Consensus of 2018 refined the Seattle Criteria, lowering that percentage further. In cases of 12-lead ECG findings that existing guidelines suggest that medical attention would be appropriate (as detailed in Table 1 below), the participant will receive a notification by text and/or email recommending that they visit their primary care physician, along with a referral letter containing the Apple Watch-derived 12-lead ECG and an outline of the findings (Table 1).

## STATISTICAL METHODS

### Sample size calculations

Sample size estimation is based on an expected cardiovascular events annual rate of 2% (at mean follow-up of 2 years) in an asymptomatic general population to detect a hazard ratio of 1.8 (A-ECG pos/neg; proportion 0.05) at α 0.05 and power 0.9, a sample size 27674 allowing for a censoring rate of 30% ^20^. A sample size of 30000 study participants would thus be adequate to meet the primary objective.

For the secondary endpoint of evaluating the correlation between changes in ECG Heart Age and activity levels, a sample size of 1000 participants is estimated. This was based on an expected correlation at r=0.15 (α 0.05, power 0.9) requiring 463 subjects and thus allowing for a combined 50% rate of dropouts to repeated ECG recording and ECG abnormalities not suitable for the estimation of ECG Heart Age.

### Analysis Plan

#### ECGs and disease prediction

Continuous variables will be described using mean and standard deviation (SD), or medians and interquartile range as appropriate. Proportional differences between groups will be assessed using the χ2 test. Comparison of group means will be performed using Student’s t test. Time-to-event data will be visually described using Kaplan-Meier event-free survival graphs. The association between smartwatch A-ECG scores and outcomes will be assessed using Cox proportional hazards regression models, unadjusted and adjusted for confounders (age, sex, BMI, smoking, hypertension, diabetes). Hazard ratios (HRs) will be presented with 95 % confidence intervals (CIs) and the assumption of proportional hazards will be confirmed using Schoenfeld’s residuals.

#### ECG Heart Age

The association of ECG Heart Age and cardiovascular outcomes will be assessed using Cox proportional hazards regression models. The correlation between ECG Heart Age and activity levels will be studied using linear regression.

#### Activity levels and cardiovascular events

The association between activity levels and cardiovascular outcomes will be assessed using Cox proportional hazards regression models, unadjusted and adjusted for confounders (age, sex and BMI).

#### Ethical considerations

The Northern Sydney Local Health District Human Research Ethics Committee approved the study (approval number 2022/ETH01269).

## Discussion

The ECG remains a foundational aspect of cardiovascular assessment, yet its acquisition requires specialized equipment operated by specialized personnel in a clinical setting. Enabling the acquisition of a fully diagnostic standard 12-lead ECG using just a smartwatch and smartphone app without any additional hardware is a first-of-its-kind opportunity, allowing clinical-grade ECG diagnostics using consumer-grade wearables.

Furthermore, conventional interpretation relies on visual waveform assessment to detect abnormalities of rate, rhythm or other pathologies such as myocardial ischemia. Early ECG changes of disease can be subtle and undiscernible to the unaided human eye ^21^. The ECG harbours a wealth of latent information that can only be extracted with advanced mathematical analyses and computational techniques ^21^. Thus, analysing SWECG using automated A-ECG could address the diagnostic yield and resource scaling limitations that thus far have precluded effective population screening with ECGs. These techniques offer superior diagnostic accuracy to the traditional 12-lead ECG analysis ^5^. This study aims to employ these recent clinical advancements and assess their real-world performance to address multiple gaps in our evidence base regarding population-level cardiovascular screening.

### A-ECG Heart Age

The A-ECG Heart Age is a prediction of an individual’s age from their ECG and is concordant with chronological age in healthy individuals but shows increasing deviation with cardiovascular risk factors and disease ^12, 13, 22^. It is a validated prognostic marker that can be intuitively communicated to the general public to encourage preventive action. This study will enable fully prospective validation of the prognostic accuracy of A-ECG Heart Age.

### Activity levels and Cardiovascular Events

CVD is substantially influenced by modifiable behavioural risk factors and physical activity is recognized as a crucial preventive measure that can reduce mortality and cardiovascular risk by 75% ^23, 24^. Despite the potential to prevent 5 million deaths annually through increased physical activity, global participation remains low with most adults failing to meet recommended levels. The reliability of self-reported activity measures often varies, prompting an increased use of activity trackers and smartwatches for more accurate assessments ^25, 26^. Studying the association between physical activity, cardiovascular risk factors, Heart Age and outcomes will provide needed evidence for future interventional preventive studies.

However, accurately quantifying physical activity in the public is challenging. Wearable devices may enable the collection of objectively measured physical activity parameters. Some cross-sectional studies report increased physical activity levels in adolescents and older adults who own such devices ^27, 28^. However, a limited number of studies report on habitual levels of activity in individuals who own an activity tracker and the associated health outcomes ^29, 30^.

Therefore, collecting physical activity data from individuals who already own a smartwatch will allow for accurate assessment of activity levels. This can be further assessed to examine the association between activity levels and the risk of cardiovascular events.

### Summary

This observational cohort study aims to evaluate the association between findings obtained from a 12-lead SWECG, combined with A-ECG analysis, in detecting cardiovascular morbidity and mortality among a large cohort of asymptomatic adults across Australia. This study explores the potential of consumer-grade technology to facilitate early detection and prevention in cardiovascular health, potentially allowing for community-based health monitoring and new preventive strategies.

### Disclosures

TTS is owner and founder of Nicollier-Schlegel SARL, which performs ECG interpretation consultancy using software that can quantify the advanced ECG measures used in the current study. TTS and MU are founders of Advanced ECG Systems, a company that is developing commercial applications of advanced ECG technology used in the current study. RK has a financial interest in Advanced ECG Systems through being married to MU.

## Data Availability

All data produced in the present study are available upon reasonable request to the authors.

## Figures

**Figure 1.**
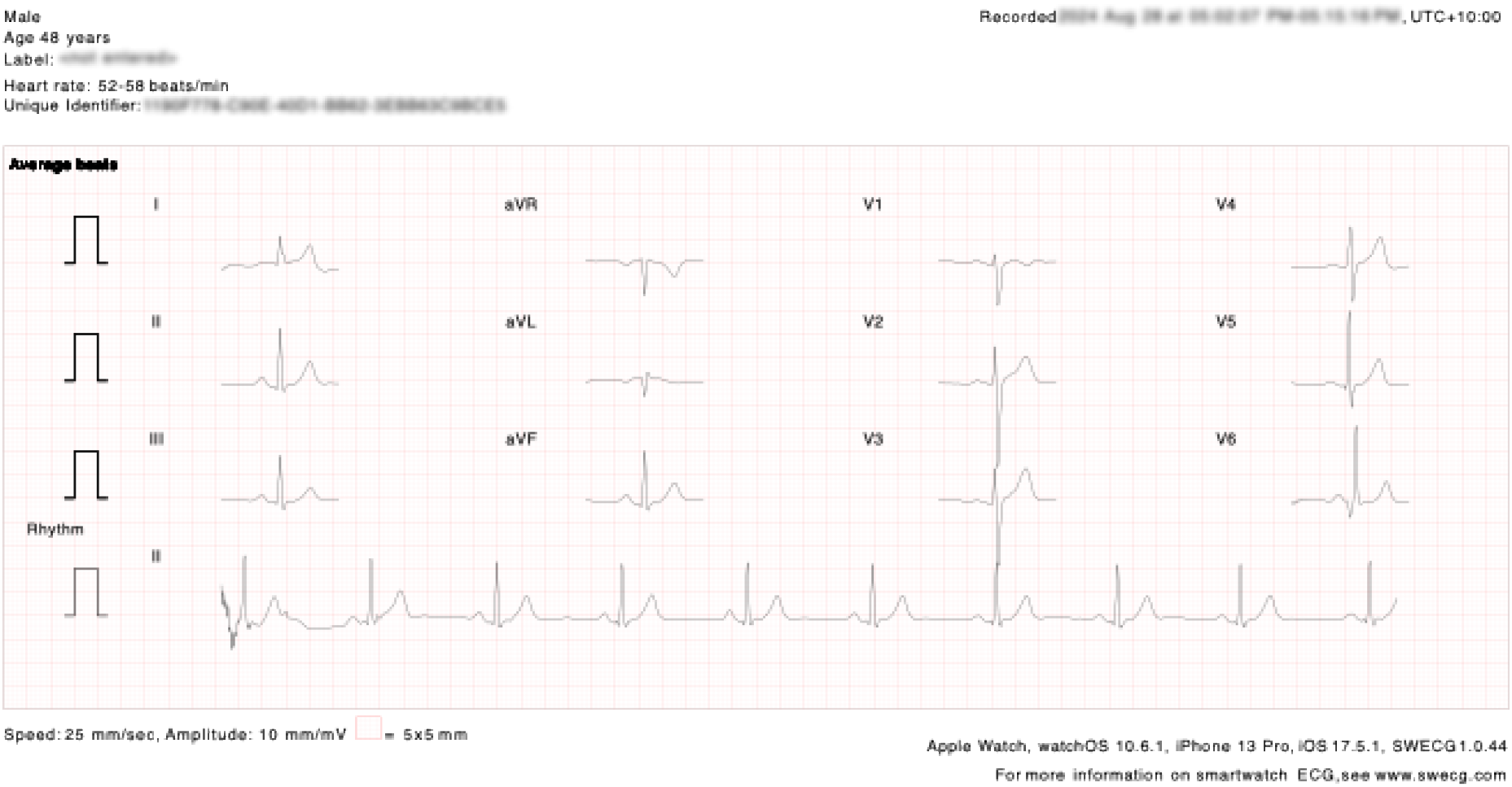
A 12-lead ECG generated using an Apple Watch and the A-ECG iPhone App

**Figure 2.**
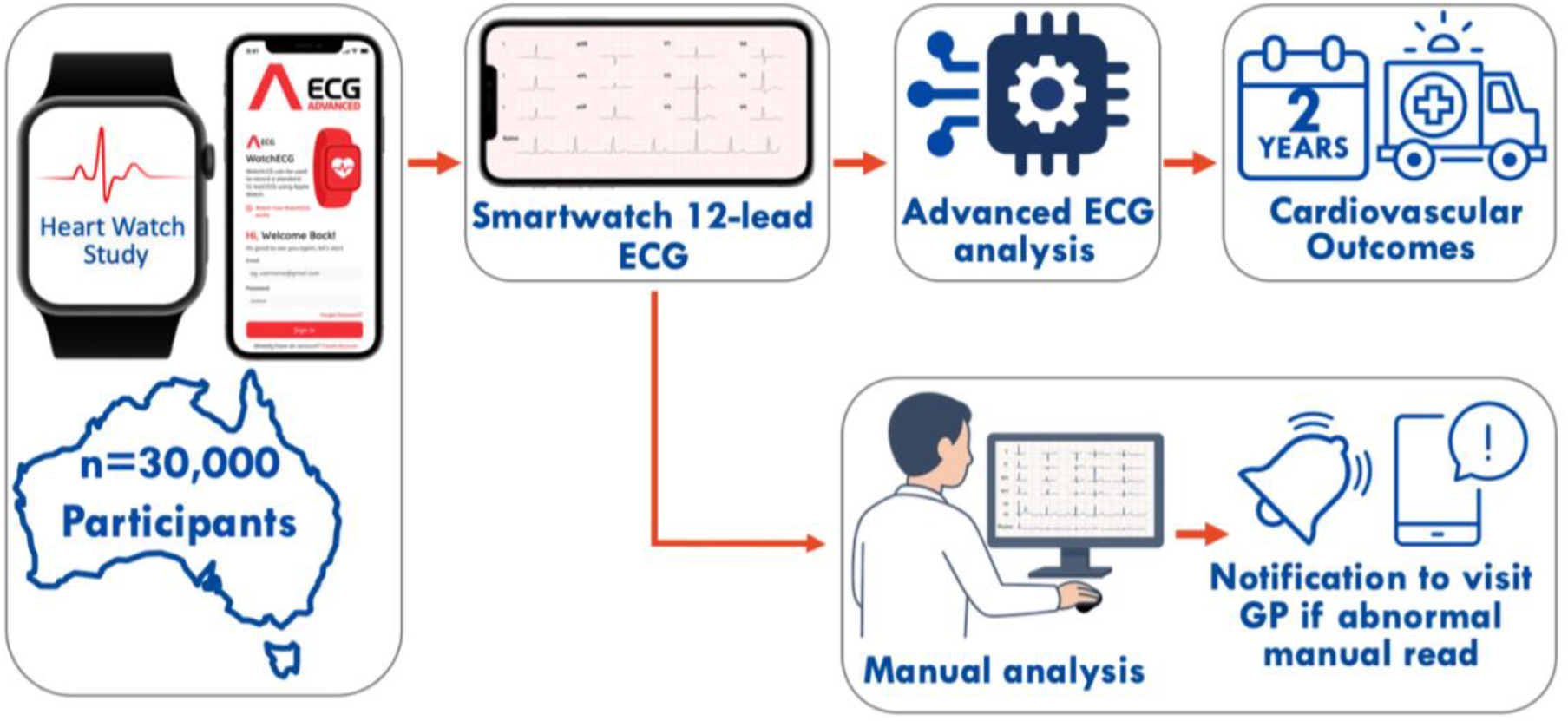
Online recruitment of 30,000 Australians and using an Apple Watch and iPhone (left panel) to generate a 12-lead Smartwatch ECG (top middle panel) then performing Advanced ECG analysis and assess cardiovascular outcomes at 2 years via data linkage (top right panels). Simultaneously, manual ECG reading is performed, and participants will be notified and given a letter to their GP if certain abnormalities are detected.

